## Supplementary methods and additional analyses for "Pneumococcal conjugate vaccine coverage and indirect protection against invasive pneumococcal disease and pneumonia hospitalisations in Australia: a retrospective record linkage cohort study"

### S1 File. Supplementary Methods and Additional Analyses

#### A. Supplementary methods

##### Table 1: Description of study covariates

| DAG variables | Study variables | Categories | Description |
|---|---|---|---|
| Age | Age group | 0-11 months, 12-23 months, 24-35 months, 36-47 months, 48-59 months | |
| Time | Year | 2001-2014 (continuous) | We adjusted for time trends using study year as a continuous variables |
| Season | Season | Winter (April-September), Summer (October-March) | |
| Socio-economic status | Index of relative socio-economic advantage and disadvantage (IRSAD) | 0-10% (most disadvantaged), 11-25%, 26-75%, 76-90%, 91-100% (least disadvantaged) | The IRSAD is one of the Socio-Economic Indexes for Areas (SEIFA) derived by the Australian Bureau of Statistics (ABS). Each index measures a different aspect of the people living in a particular area and ranks the different geographical areas across Australia according to a score based on the socio-economic characteristics in that area. The IRSAD score is derived from 21 different variables which include low or high income, internet connection, skilled or unskilled occupations and education. The IRSAD scores included in our analyses were based on the mother's residential address at the time of her child's birth. |
| Remoteness | Accessibility/Remoteness Index of Australia (ARIA) | Major cities, inner regional, outer regional, remote, very remote | ARIA is a standard national measure of geographic remoteness and access to services for localities and areas throughout Australia and is classified into major cities, inner regional, outer regional, remove and very remote. Like the IRSAD, the ARIA classification used in this study was based on the mother's residential address at the time of her child's birth |
| Low birth weight | Birth weight | <1500 grams, 1500-2499 grams, 2500-3499 grams, 3500-4499 grams and ≥ 4500 grams | Proportion of optimal birth weight (POBW) was used rather than stand-alone birth weight. This is a calculated measure of the appropriateness of intrauterine growth which takes into account gestational age, maternal age, maternal height, parity and infant gender. [1] |

| | | | |
|---|---|---|---|
| Prematurity | Gestational age | <33 weeks | |
| Parental smoking | Maternal smoking during pregnancy | Yes/No | |
| Siblings | Number of previous pregnancies | 0, 1 and ≥2 | |
| High risk comorbidity | Any hospitalisation < 6 weeks after birth | Yes/No | |
| | Any hospital admission for category A risk condition | Yes/No | Primary of secondary ICD-10-AM diagnosis code for a comorbid condition associated with highest increased risk of IPD recorded before IPD or pneumonia onset. [2]<br><br>**<u>Category A conditions associated with the highest risk of invasive pneumococcal disease:</u>**<br>Functional or anatomical asplenia, including:<br>• Sickle cell disease or other haemoglobinopathies<br>• Congenital or acquired asplenia (for example, splenectomy) or splenic dysfunction<br>Immunocompromising conditions, including:<br>• Congenital or acquired immune deficiency including symptomatic IgG subclass or isolated IgA deficiency<br>• Immunosuppressive therapy (including corticosteroid therapy ≥ 2mg per kg per day or prednisolone or equivalent for more than 1 week) or ration therapy, where there is sufficient immune reconstitution for vaccine response to be expected<br>Haematological and other malignancies<br>Solid organ transplant<br>Haematopoietic stem cell transplant<br>HIV infection (including AIDS)<br>Chronic renal failure or relapsing or persistent nephrotic syndrome<br>Proven or presumptive cerebrospinal fluid leak<br>Cochlear implants<br>Intracranial shunts |
| | Any hospital admission for category B risk condition | Yes/No | Primary of secondary ICD-10-AM diagnosis code for a comorbid condition associated increased risk of IPD recorded before IPD or pneumonia onset. [2]<br><br>**<u>Category B conditions associated with increased risk of invasive pneumococcal disease:</u>**<br>Chronic cardiac disease:<br>• Cyanotic heart disease or cardiac failure in children<br>• Excluding hypertension only (in adults)<br>Chronic lung disease<br>• Chronic lung disease in preterm infants<br>• Cystic fibrosis<br>Severe asthma in adults<br>Diabetes<br>Down syndrome<br>Alcoholism<br>Chronic liver disease<br>Preterm birth at <28 weeks gestation<br>Tobacco smoking<br>History of previous invasive pneumococcal disease in children |
| Influenza vaccination | Annual influenza vaccination | Yes/No (for each year) | At least one dose of influenza vaccine received that year |
| Indigenous | Indigenous status | Indigenous and non-Indigenous | Indigenous status was derived using three algorithms pro posed by Christensen et al[3], which provides an application of the theory |

| | | | |
|---|---|---|---|
| | | | outlined in 'National Best Practice Guidelines for Data Linkage Activities Relating to Aboriginal and Torres Strait Islander People' |

**Table 2: 10th Revision of the International Classification of Diseases Australian Modification (ICD-10-AM) codes for classification of pneumonia hospitalisation**

| ICD-10-AM code | Description |
|---|---|
| J13 | Pneumococcal pneumonia |
| J18.1 | Lobar pneumonia, unspecified |
| J10.0 | Influenza with pneumonia, influenza virus identified |
| J11.0 | Influenza with pneumonia, virus not identified |
| J12.0 | Pneumonia due to adenovirus |
| J12.1 | Pneumonia due to respiratory syncytial virus |
| J12.2 | Pneumonia due to parainfluenza virus |
| J12.8 | Pneumonia due to other virus not elsewhere classified |
| J12.9 | Viral pneumonia, unspecified |
| J15.0 | Pneumonia due to Klebsiella pneumoniae |
| J15.1 | Pneumonia due to Pseudomonas |
| J15.2 | Pneumonia due to Staphylococcus |
| J15.3 | Pneumonia due to Streptococcus, group B |
| J15.4 | Pneumonia due to other Streptococci |
| J15.5 | Pneumonia due to Escherichia coli |
| J15.6 | Pneumonia due to other aerobic Gram-negative bacteria |
| J15.8 | Other bacterial pneumonia |
| J15.7 | Pneumonia due to Mycoplasma pneumoniae |
| J15.9 | Bacterial pneumonia, unspecified |
| J16.0 | Chlamydial pneumonia |
| J16.8 | Pneumonia due to other specified infectious organisms |
| J17.0 | Pneumonia in bacterial diseases classified elsewhere |
| J17.1 | Pneumonia in viral diseases classified elsewhere |
| J17.2 | Pneumonia in mycoses |
| J17.3 | Pneumonia in parasitic diseases |
| J17.8 | Pneumonia in other infectious diseases classified elsewhere |
| B01.2 | Varicella pneumonia |
| B05.2 | Measles complicated by pneumonia |
| B37.1 | Pulmonary candidiasis |
| B59 | Pneumocystosis |
| J18.0 | Bronchopneumonia, unspecified |
| J18.2 | Hypostatic pneumonia, unspecified |
| J18.8 | Other pneumonia, organism unspecified |
| J18.9 | Pneumonia, organism unspecified |

**Table 3: Characteristics of children who are under-vaccinated against any pneumococcal conjugate vaccine by year, New South Wales and Western Australia, 2001-2012**

| | Low birth weight | Preterm | Maternal parity >1 | Maternal smoking | Indigenous | High-risk hospitalisation* | Any hospitalisation < 6 weeks |
|---|---|---|---|---|---|---|---|
| | n (%) | n (%) | n (%) | n (%) | n (%) | n (%) | n (%) |
| **2001** | 4 599 (4·4) | 763 (0·7) | 29 204 (28·0) | 18 321 (17·5) | 4 754 (4·6) | 3 450 (3·3) | 13 046 (12·5) |

| | | | | | | | |
|---|---|---|---|---|---|---|---|
| **2002** | 9 098 (4·4) | 1 403 (0·7) | 57 988 (27·8) | 35 628 (17·1) | 9 461 (4·5) | 7 478 (3·6) | 28 471 (13·6) |
| **2003** | 13 533 (4·3) | 2 036 (0·7) | 86 245 (27·5) | 51 702 (16·5) | 14 187 (4·5) | 11 481 (3·7) | 43 589 (13·9) |
| **2004** | 18 103 (4·3) | 2 727 (0·7) | 114 340 (27·4) | 67 067 (16·1) | 19 006 (4·6) | 15 329 (3·7) | 59 213 (14·2) |
| **2005** | 22 815 (4·3) | 3 423 (0·7) | 176 464 (27·5) | 82 909 (15·7) | 24 254 (4·6) | 19 437 (3·7) | 76 144 (14·4) |
| **2006** | 27 714 (4·3) | 4 172 (0·7) | 180 074 (27·4) | 98 353 (15·4) | 29 818 (4·6) | 23 592 (3·7) | 92 881 (14·5) |
| **2007** | 28 333 (4·3) | 4 289 (0·7) | 184 796 (27·4) | 95 649 (14·6) | 31 003 (4·7) | 24 588 (3·7) | 97 838 (14·9) |
| **2008** | 29 483 (4·3) | 4 443 (0·7) | 189 364 (27·5) | 93 656 (14·0) | 32 322 (4·8) | 25 010 (3·7) | 100 835 (15·0) |
| **2009** | 29 974 (4·3) | 4 574 (0·7) | 193 610 (27·5) | 92 198 (13·4) | 33 747 (4·9) | 25 260 (3·7) | 103 956 (15·1) |
| **2010** | 30 386 (4·2) | 4 645 (0·7) | 194 925 (27·3) | 89 789 (12·8) | 35 159 (5·0) | 25 776 (3·7) | 106 976 (15·2) |
| **2011** | 30 400 (4·2) | 4 689 (0·7) | 193·925 (27·0) | 86 648 (12·1) | 36 081 (5·0) | 26 066 (3·6) | 109 443 (15·3) |
| **2012** | 25 217 (4·2) | 3 839 (0·6) | 161 171 (26·8) | 82 427 (11·5) | 36 335 ((5·0) | 26 309 (3·7) | 112 773 (15·7) |

* See supplementary Table 2 for fill list of high-risk comorbidities

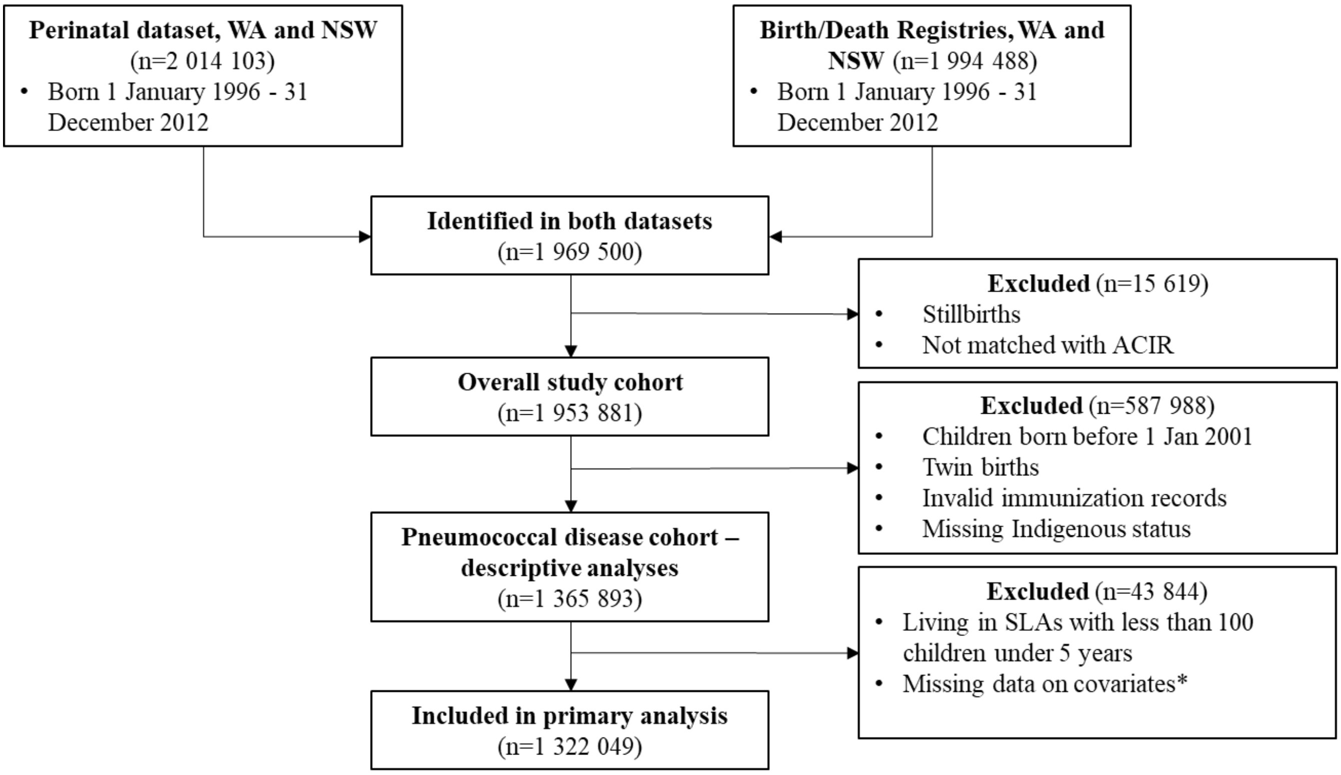

* Covariates: statistical local area (SLA), index of relative socio-economic advantage and disadvantage (IRSAD), maternal age at delivery, accessibility/remoteness index of Australia (ARIA), birth weight, gestational age, maternal smoking during pregnancy, number of previous pregnancies, hospitalisations for high-risk medical comorbidities

**Fig 1: Flow chart of study cohort, including data sources**

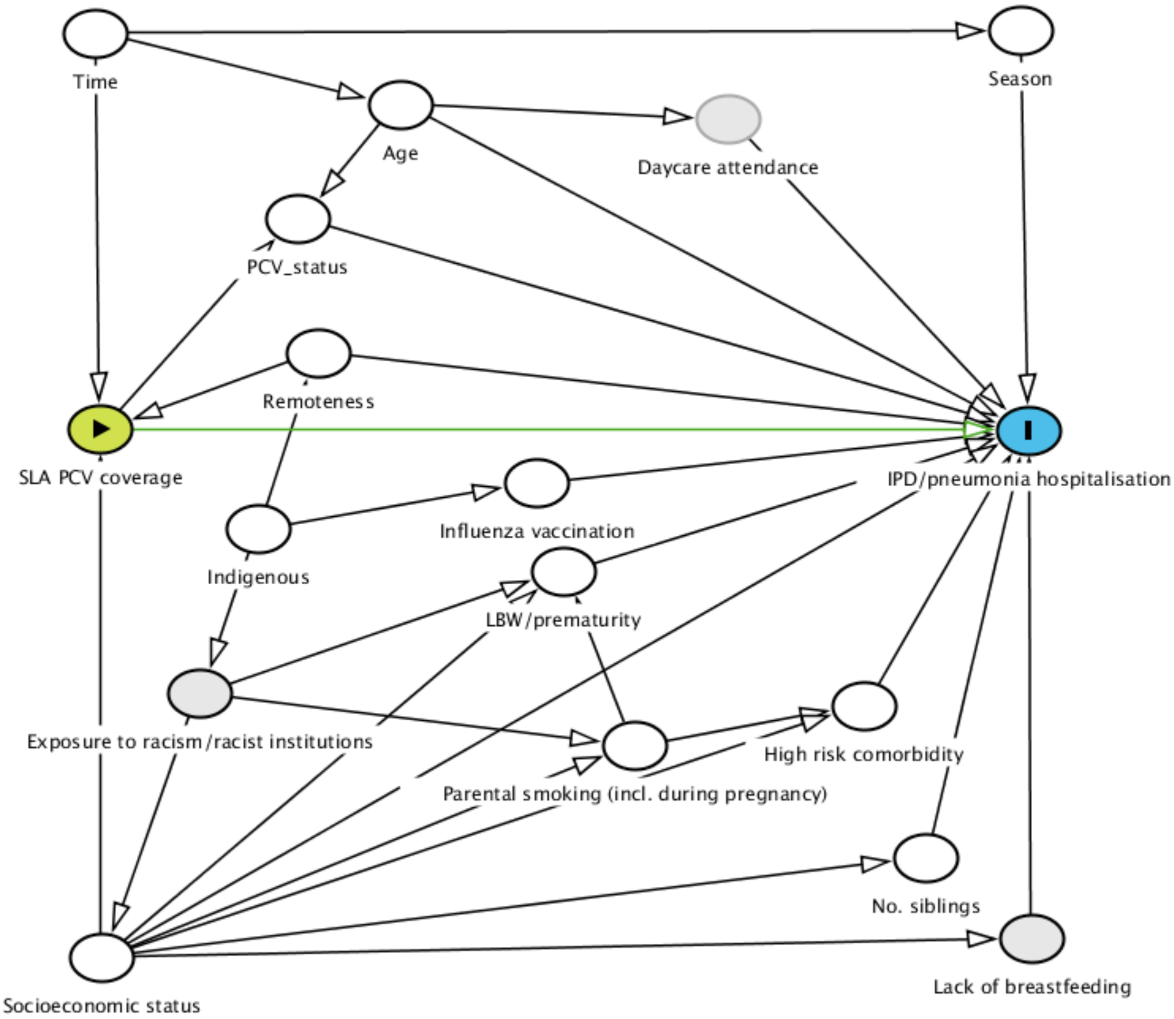

**Fig 2: Directed acyclic graph**

Fig 2 is a directed acyclic graph (DAG).[4] Arrows are used to indicate causal pathways between exposure (PCV coverage) and outcome (IPD or pneumonia hospitalisation among under-vaccinated children). The green line highlights the causal relationship under investigation. The diagram includes both measured (dark grey) and unmeasured (light grey) covariates. Since our analysis includes only under-vaccinated person-time, we have represented this as conditioning on PCV status in the DAG above.We identified the following potential confounders for adjustment (white): age, season, influenza vaccination, socioeconomic status, remoteness, low birth weight, prematurity, parental smoking, number of siblings and high risk comorbidity. The study variables that we use as proxies for the potential confounders identified are listed in Table 2. In most cases, there was a direct correlation between potential variables listed in the DAG and study variables used for adjustment, with the exception of number of siblings – for which we used parity as a proxy since this variable was not available. A table of the trends in key confounders over time is presented below (Table 10). Child's influenza vaccination status was also initially investigated but excluded from final models due to small numbers of recorded doses and collinearity with PCV. Despite having unmeasured potential confounders including daycare attendance, breastfeeding and exposure to racism and racist institutions, our diagram suggests that the relationship being examined, between SLA-level PCV coverage and risk of IPD or pneumonia hospitalisation, is unlikely to be affected by the omission of those factors.

#### B. Incidence of invasive pneumococcal disease

##### Table 4: Rate of invasive pneumococcal disease (IPD) among all children and under-vaccinated children by vaccination period age group and Indigenous status, New South Wales and Western Australia, 2001-2013

| Age group | PCV period | All non-Indigenous children | | Under-vaccinated* non-Indigenous children | | All Indigenous children | | Under-vaccinated* Indigenous children | |
|---|---|---|---|---|---|---|---|---|---|
| | | Cases per person-years | IPD rate (95% CI) † | Cases per person-years | IPD rate (95% CI)† | Cases per person-years | IPD rate (95% CI) † | Cases per person-years | IPD rate (95% CI) † |
| **PCV7-type IPD** | | | | | | | | | |
| 0-11m | Pre-universal PCV (2001-04) | 159/349 502 | 45·5 (38·9-53·1) | 159/ 346 727 | 45·9 (39·2-53·6) | 8/16 613 | 48·2 (24·1-96·3) | 8/13 578 | 58·9 (29·5-117·8) |
| | Post-PCV7 (2005-10) | 27/663 322 | 4·1 (2·8-5·9) | 21/285 657 | 7·4 (4·8-11·3) | <5/34 487 | 8·7 (2·8-27·0) | <5/16 704 | 18·0 (5·8-55·7) |
| | Post-PCV13 (2011-12) | 2/284 374 | 0·7 (0·2-2·8) | 2/101 319 | 2·0 (0·5-7·9) | <5/15 125 | 6·6 (0·9-46·9) | <5/5 973 | 16·9 (2·4-118·8) |
| 12-23 m | Pre-universal PCV (2001-04) | 221/248 721 | 88·9 (77·9-101·4) | 219/243 604 | 90·0 (78·7-102·6) | 5/11 783 | 42·4 (17·7-101·9) | 5/7 059 | 70·8 (29·5-170·2) |
| | Post-PCV7 (2005-10) | 46/647 801 | 7·1 (5·3-9·5) | 29/70 951 | 40·9 (28·4-58·8) | <5/33 030 | 3·0 (0·4-21·5) | 0/4 160 | 0 |
| | Post-PCV13 (2011-12) | 2/346 006 | 0·6 (0·1-2·3) | 0/17 888 | 0 | 0/18 627 | 0 | 0/1 252 | 0 |
| 24-59m | Pre-universal PCV (2001-04) | 65/199 202 | 32·6 (25·6-41·6) | 63/1955 155 | 32·3 (25·1-41·3) | <5/9 511 | 21·0 (5·3-84·1) | <5/5 795 | 17·5 (2·5-124·1) |
| | Post-PCV7 (2005-10) | 67/1 809 221 | 3·7 (2·9-4·7) | 50/547 895 | 9·1 (6·9-12·0) | 0/88 812 | 0 | 0/19 651 | 0 |
| | Post-PCV13 (2011-12) | 7/1 023 209 | 0·7 (0·3-1·4) | 0/45 384 | 0 | <5/54 480 | 1·8 (0·3-13·0) | 0/3 273 | 0 |
| **PCV13, non-PCV7-type IPD** | | | | | | | | | |
| 0-11m | Pre-universal PCV (2001-04) | 11/349 543 | 3·1 (1·7-5·7) | 11/349 543 | 3·1 (1·7-5·7) | <5/16 616 | 6·0 (0·8-42·7) | <5/16 616 | 6·0 (0·8-42·7) |
| | Post-PCV7 (2005-10) | 79/663 311 | 11·9 (9·6-14·8) | 79/663 263 | 11·9 (9·6-14·8) | 8/34 486 | 23·7 (11·8-47·3) | 8/34 485 | 23·7 (11·8-47·3) |
| | Post-PCV13 (2011-12) | 19/284 366 | 6·7 (4·3-10·5) | 18/174 742 | 10·3 (6·5-16·3) | <5/15 125 | 19·8 (6·4-61·5) | <5/9 527 | 31·5 (10·2-97·6) |
| 12-23m | Pre-universal PCV (2001-04) | 9/248 937 | 3·6 (1·9-6·9) | 9/248 937 | 3·6 (1·9-6·9) | <5/11 791 | 8·4 (1·2-60·2) | <5/11 791 | 8·4 (1·2-60·2) |
| | Post-PCV7 (2005-10) | 81/647 809 | 12·5 (10·1-15·5) | 81/647 760 | 12·5 (10·1-15·5) | 9/33 022 | 27·2 (14·1-52·4) | 9/33 020 | 27·3 (14·2-52·4) |
| | Post-PCV13 (2011-12) | 32/345 959 | 9·2 (6·5-13·1) | 25/190 621 | 13·1 (8·9-19·4) | <5/18 623 | 16·1 (5·2-49·9) | <5/10 096 | 29·7 (9·6-92·1) |
| 24-59m | Pre-universal PCV (2001-04) | 6/199 457 | 3·0 (1·4-6·7) | 6/199 457 | 3·0 (1·4-6·7) | <5/9 523 | 10·5 (1·5-74·5) | <5/9 523 | 10·5 (1·5-74·5) |
| | Post-PCV7 (2005-10) | 6/199 457 | 3·0 (1·4-6·7) | 6/199 457 | 3·0 (1·4-6·7) | <5/9 523 | 10·5 (1·5-74·5) | <5/9 523 | 10·5 (1·5-74·5) |
| | Post-PCV13 (2011-12) | 47/1 023 471 | 4·7 (3·5-6·2) | 46/850 646 | 5·4 (4·1-7·2) | 6/54 440 | 11·0 (5·0-24·5) | 5/46 391 | 12·9 (5·8-28·8) |
| **All-cause IPD** | | | | | | | | | |
| 0-11m | Pre-universal PCV (2001-04) | 222/349 481 | 63·5 (55·7-72·4) | 222/346 706 | 64·0 (56·1-73·0) | 14/16 611 | 84·3 (49·9-142·3) | 14/13 577 | 103·1 (61·1-174·1) |
| | Post-PCV7 (2005-10) | 151/663 278 | 22·8 (55·7-72·4) | 56/285 648 | 19·6 (15·1-25·5) | 20/34 480 | 58·0 (37·4-89·9) | 8/16 703 | 47·9 (24·0-95·8) |
| | Post-PCV13 (2011-12) | 41/284 356 | 14·4 (10·6-19·6) | 14/ 101 316 | 13·8 (8·2-23·3) | 8/15 122 | 52·9 (26·5-105·8) | <5/5 973 | 67·0 (25·1-178·4) |
| 12-23m | Pre-universal PCV (2001-04) | 274/248 645 | 110·2 (97·9-124·0) | 272/243 536 | 111·7 (99·2-125·8) | 9/11 777 | 76·4 (39·7-146·9) | 7/7 056 | 99·2 (47·3-208·1) |
| | Post-PCV7 (2005-10) | 171/647 631 | 26·4 (22·7-30·7) | 40/70 934 | 57·8 (42·6-78·5) | 16/33 005 | 51·5 (32·0-82·9) | 0/4 159 | 0 |
| | Post-PCV13 (2011-12) | 67/345 913 | 19·4 (15·2-24·6) | <5/17 883 | 16·8 (5·4-55·0) | 6/18 618 | 32·2 (14·5-71·7) | <5/1 251 | 79·9(11·3-567·3) |
| 24-59m | Pre-universal PCV (2001-04) | 80/199 108 | 40·2 (32·3-50·0) | 78/195 072 | 40·0 (32·0-49·9) | <5/9 504 | 42·1 (15·8-112·1) | <5/5 791 | 34·5 (8·6-138·1) |
| | Post-PCV7 (2005-10) | 219/1 808 359 | 12·1 (10·6-13·8) | 73/547 666 | 13·3 (10·6-16·8) | 17/88 711 | 19·2 (11·9-30·8) | <5/19 640 | 15·3 (4·9-47·4) |
| | Post-PCV13 (2011-12) | 94/1 023 229 | 9·2 (7·5-11·2) | 8/45 353 | 17·6 (8·8-35·3) | 12/53 493 | 23·9 (13·8-41·2) | 0/ 3 268 | 0 |

*Under-vaccinated against any PCV for PCV7-type and all-cause IPD results; under-vaccinated against PCV13 for PCV13, non-PCV7-type IPD results; †Per 100 000 person-years

#### C. Incidence of pneumonia hospitalisations

**Table 5: Annual incidence of pneumonia hospitalisation among all children and under-vaccinated children (any PCV) by vaccination period, age group and Indigenous status, New South Wales and Western Australia, 2001-2013**

| | | All non-Indigenous children | | Under-vaccinated non-Indigenous children* | | All Indigenous children | | Under-vaccinated Indigenous children* ( | |
|---|---|---|---|---|---|---|---|---|---|
| | | Cases per person-years | Hospitalisation rate (CI) † | Cases per person-years | Hospitalisation rate (CI) † | Cases per person-years | Hospitalisation rate (CI) † | Cases per person-years | Hospitalisation rate (CI) † |
| **All-cause pneumonia** | | | | | | | | | |
| 0-11m | Pre-universal PCV (2001-04) | 2 493/335 652 | 74·3 (71·4-77·2) | 2 468/332 740 | 74·2 (71·3-77·2) | 591/15 800 | 374·1 (345·1-405·5) | 448/12 774 | 342·8 (312·2-376·5) |
| | Post-PCV7 (2005-10) | 3 424/661 708 | 51·7 (50·0-53·5) | 1 395/284 955 | 49·0 (46·5-51·6) | 751/34 169 | 219·8 (204·6-236·1) | 410/16 513 | 248·3 (225·4-273·6) |
| | Post-PCV13 (2011-12) | 1 486/284 134 | 52·3 (49·7-55·0) | 525 /101 296 | 51·8 (47·6-56·5) | 259/15 031 | 172·6 (152·8-195·0) | 109/5 946 | 183·3 (151·9-221·1) |
| 12-23 m | Pre-universal PCV (2001-04) | 2 746/245 578 | 111·8 (107·7-116·1) | 2 674/240 540 | 111·2 (107·0-115·5) | 338/11 347 | 297·9 (267·7-331·4) | 156/6 854 | 227·6 (194·5-266·3) |
| | Post-PCV7 (2005-10) | 4 763/642 403 | 74·1 (72·1-76·3) | 479/70 682 | 67·8 (62·0-74·1) | 654/32 114 | 203·6 (188·6-219·9) | 77/4 042 | 190·5 (152·4-238·2) |
| | Post-PCV13 (2011-12) | 2 473/343 684 | 72·0 (69·2-74·8) | 73 /17 800 | 41·0 (32·6-51·6) | 303/18 225 | 166·3 (148·5-186·1) | 28/1 222 | 229·1 (158·2-331·8) |
| 24-59m | Pre-universal PCV (2001-04) | 1 381/194 400 | 71·0 (67·4-74·9) | 1 348/190 511 | 70·8 (67·1-74·6) | 155/8 982 | 172·6 (147·4-202·0) | 72/5 537 | 130·0 (103·2-163·8) |
| | Post-PCV7 (2005-10) | 7 708/1 774 448 | 43·4 (42·5-44·4) | 2 076/536 443 | 38·7 (37·1-40·4) | 726/84 222 | 86·2 (80·2-92·7) | 152/18 703 | 81·3 (69·3-95·3) |
| | Post-PCV13 (2011-12) | 4 102/1 007 430 | 40·7 (39·5-42·0) | 123/44 880 | 27·4 (23·0-32·7) | 386/52 201 | 73·9 (66·9-81·7) | 18/3 111 | 57·9 (36·5-91·8) |
| **Pneumococcal and lobar pneumonia** | | | | | | | | | |
| 0-11m | Pre-universal PCV (2001-04) | 87/336 715 | 2·6 (2·1-3·2) | 87/333 957 | 2·6 (2·1-3·2) | 19/15 971 | 11·9 (7·6-18·7) | 15/12 944 | 11·6 (7·0-19·2) |
| | Post-PCV7 (2005-10) | 66/663 244 | 1·0 (0·8-1·3) | 27/285 878 | 0·9 (0·6-1·4) | 16/34 475 | 4·6 (2·8-7·6) | 8/16 706 | 4·8 (2·4-9·6) |
| | Post-PCV13 (2011-12) | 23/284 872 | 0·8 (0·5-1·2) | 6/101 519 | 0·6 (0·3-1·3) | 7/15 151 | 4·6 (2·2-9·7) | <5/5 986 | 6·7 (2·5-17·8) |
| 12-23 m | Pre-universal PCV (2001-04) | 117/248 378 | 4·7 (3·9-5·6) | 114/243 277 | 4·7 (3·9-5·6) | 13/ 11 748 | 11·1 (6·4-19·1) | 8/7 045 | 11·4 (5·7-22·7) |
| | Post-PCV7 (2005-10) | 92/647 712 | 1·4 (1·2-1·7) | 17/71 346 | 2·4 (1·5-3·8) | 24/32 996 | 7·3 (4·9-10·9) | <5/4 164 | 9·6 (3·6-25·6) |
| | Post-PCV13 (2011-12) | 39/346 473 | 1·1 (0·8-1·5) | 2/17 911 | 1·1 (0·3-4·5) | 10/18 643 | 5·4 (2·9-10·0) | <5/1 254 | 15·9 (4·0-63·8) |
| 24-59m | Pre-universal PCV (2001-04) | 41/198 431 | 2·1 (1·5-2·8) | 38/194 398 | 2·0 (1·4-2·7) | 7/9 453 | 7·4 (3·5-15·5) | <5/5 772 | 5·2 (1·7-16·1) |
| | Post-PCV7 (2005-10) | 146/1 808 890 | 0·8 (0·7-0·9) | 42/548 937 | 0·8 (0·6-1·0) | 20/88 622 | 2·3 (1·5-3·5) | <5/19 641 | 1·5 (0·5-4·7) |
| | Post-PCV13 (2011-12) | 66/1 025 012 | 0·6 (0·5-0·8) | 4/45 433 | 0·9 (0·3-2·3) | 13/54 489 | 2·4 (1·4-4·1) | 0/3 278 | 0 |

*Under-vaccinated against any PCV; †Per 10 000 person years

#### D. Incidence of invasive pneumococcal disease and vaccine coverage

**Table 6: Annual pneumococcal conjugate vaccine coverage (PCV) and rate of 7-valent PCV-type invasive pneumococcal disease (IPD) among all children and under-vaccinated children and among children under five years of age, New South Wales and Western Australia, 2005-2012**

| Year | All children | | Under-vaccinated children | | Any PCV coverage, median (IQR) |
|---|---|---|---|---|---|
| | Cases/person-years | PCV7-type IPD rate | Cases/person years | PCV7-type IPD rate | |
| **2001** | 3/52 599 | 0.6 (0.2-1.8) | <5/52 537 | 0.6 (0.2-1.8) | - |
| **2002** | 98/ 156 546 | 6.3 (5.1-7.6) | 98/154 962 | 6.3 (5.2-7.7) | - |
| **2003** | 157/ 260 627 | 6.0 (5.2-7.0) | 155/254 963 | 6.1 (5.2-7.1) | - |
| **2004** | 202/365 561 | 5.3 (4.8-6.3) | 199/349 455 | 5.7 (5.0-6.5) | - |
| **2005** | 82/472 529 | 1.7 (1.4-2.2) | 76/322 142 | 2.4 (1.9-3.0) | 17·5 (15·8-19·2) |
| **2006** | 25/532 325 | 0.5 (0.3-0.7) | 16/238 373 | 0.7 (0.4-1.1) | 27·2 (24·7-29·2) |
| **2007** | 14/545 472 | 0.3 (0.2-0.4) | <5/150 083 | 0.3 (0.1-0.7) | 38·1 (35·6-40·3) |
| **2008** | 12/560 578 | 0.2 (0.1-0.4) | <5/90 950 | 0.3 (0.1-1.0) | 46·1 (43·0-48·1) |
| **2009** | 7/575 653 | 0.1 (0.1-0.3) | <5/72 796 | 0.4 (0.1-1.3) | 53·5 (50·7-55·9) |
| **2010** | 4/590 098 | 0.1 (0.0-0.2) | <5/70 675 | 0.1 (0.0-1.0) | 62·0 (59·1-63·9) |
| **2011** | 5/598 864 | 0.1 (0.0-0.2) | <5/70 602 | 0.3 (0.1-1.1) | 72·4 (69·7-74·5) |
| **2012** | 5/602 610 | 0.1 (0.0-0.2) | 0/71 514 | 0.0 | 86·8 (84·5-88·9) |

*Calculated as of 31 December of each year

**Table 7: Annual 13-valent pneumococcal conjugate vaccine coverage (PCV13) and rate of PCV13, non PCV7-type invasive pneumococcal disease (IPD) among all children and under-vaccinated children and among children under five years of age, New South Wales and Western Australia, 2005-2012**

| Year | All children | | Under-vaccinated children | | PCV13 coverage, median (IQR) |
|---|---|---|---|---|---|
| | Cases/person-years | PCV7-type IPD rate | Cases/person years | PCV7-type IPD rate | |
| **2001** | 0/52 600 | 0.0 | 0/52 600 | 0.0 | - |
| **2002** | 5/156 588 | 0.3 (0.1-0.8) | 5/156 588 | 0.3 (0.1-0.8) | - |
| **2003** | 6/260 790 | 0.2 (0.1-0.5) | 6/260 791 | 0.2 (0.1-0.5) | - |
| **2004** | 18/365 890 | 0.5 (0.3-0.8) | 18/365 890 | 0.5 (0.3-0.8) | - |
| **2005** | 29/472 984 | 0.6 (0.4-0.9) | 29/472 984 | 0.6 (0.4-0.9) | - |
| **2006** | 19/532 730 | 0.4 (0.2-0.6) | 19/532 730 | 0.4 (0.2-0.6) | - |
| **2007** | 38/545 702 | 0.7 (0.5-1.0) | 38/545 700 | 0.7 (0.5-1.0) | - |
| **2008** | 59/560 632 | 1.0 (0.8-1.4) | 59/560 622 | 1.1 (0.8-1.4) | - |
| **2009** | 54/575 567 | 0.9 (0.7-1.2) | 54/575 539 | 0.9 (0.7-1.2) | - |
| **2010** | 66/589 960 | 1.1 (0.9-1.4) | 66/589 869 | 1.1 (0.9-1.4) | - |
| **2011** | 62/598 699 | 1.0 (0.8-1.3) | 62/584 674 | 1.1 (0.8-1.4) | 6·8 (5·6-8·1) |
| **2012** | 32/602 458 | 0.5 (0.4-0.8) | 28/430 478 | 0.7 (0.4-0.9) | 32·1 (29·1-35·4) |

#### E. Descriptive analyses of coverage and indirect protection against invasive pneumococcal disease

**Table 8: Crude rates of invasive pneumococcal disease (IPD) among under-vaccinated children by level of PCV coverage within their statistical local area of residence, New South Wales and Western Australia, 2001-2012**

| | PCV coverage category, % | Cases per Person-Years | IPD rate (95% CI)* |
|---|---|---|---|
| **PCV7-type IPD** | <25 | 459/856 101 | 53·6 (48·9-58·8) |
| | 25-49 | 60/279 606 | 21·5 (16·7-27·6) |
| | 50-74 | 18/297 908 | 6·0 (3·8-9·6) |
| | 75-100 | 9/389 787 | 2·3 (1·2-4·4) |
| **PCV13, non-PCV7-type IPD** | <25 | 181/1 791 802 | 10·1 (8·7-11·7) |
| | 25-49 | 22/300 608 | 7·3 (4·8-11·1) |
| | 50-74 | 0/336 | 0 |
| | 75-100 | - | - |

**Table 9: Rate of all-cause pneumonia hospitalisation among under-vaccinated children by level of PCV coverage within their statistical local area of residence, New South Wales and Western Australia, 2001-2012**

| | PCV coverage category, % | Cases/person-years | Hospitalisation rate (95% CI)* |
|---|---|---|---|
| **All-cause pneumonia hospitalisation** | <25 | 8 010/977 306 | 82·0 (80·2-83·8) |
| | 25-49 | 1 044/239 797 | 43·5 (41·0-46·3) |
| | 50-74 | 371/230 648 | 42·1 (39·5-44·8) |
| | 75-100 | 2 010/386 541 | 53·8 (51·4-56·4) |
| **Pneumococcal and lobar pneumonia hospitalisation** | <25 | 282/989 338 | 2·9 (2·5-3·2) |
| | 25-49 | 26/244 885 | 1·0 (0·7-1·6) |
| | 50-74 | 15/234 692 | 0·6 (0·4-1·1) |
| | 75-100 | 23/335 652 | 0·7 (0·5-1·0) |

*Per 10 000 person years

#### F. Association between coverage and indirection protection against pneumococcal disease

**Table 10: Crude and adjusted* incidence rate ratios of 7-valent pneumococcal conjugate vaccine-type (PCV7-type) invasive pneumococcal disease (IPD) among under-vaccinated children under five years, by percent increase in pneumococcal conjugate vaccine (PCV) coverage† among children under five years of age, New South Wales and Western Australia, 2009-2012**

| | **Crude** | | **Adjusted*** | |
|---|---|---|---|---|
| | Incidence rate ratio (95% CI) | P-value | Incidence rate ratio (95% CI) | P-value |
| **PCV7-type invasive pneumococcal disease (n= 8 579 771§)** | 0.965 (0.961-0.969) | <0.001 | 0.967 (0.958-0.975) | <0.001 |
| **PCV13, non-PCV7-type invasive pneumococcal disease (IPD) (n=8 941 562§)** | 0.985 (0.972-0.998) | 0.029 | 0.989 (0.971-1.007) | 0.241 |

*adjusted by Indigenous status, season, Index of Relative Socio-economic Advantage and Disadvantage (IRSAD) score, Accessibility or Remoteness Index of Australia (ARIA) category, birth weight, gestational age, maternal smoking during pregnancy, number of previous pregnancies, and previous hospitalisation for a range of high-risk medical comorbidities; †Coverage of any PCV – inclusive of both PCV7 and PCV13; ‡Akaike information criterion § Number of observations for adjusted analysis (quarterly person-time)

**Table 11: Crude and adjusted* incidence rate ratios of pneumonia hospitalisations among under-vaccinated children under five years, by percent increase in pneumococcal conjugate vaccine (PCV) coverage† among children under five years of age, New South Wales and Western Australia, 2009-2012**

| | Crude | | Adjusted | |
|---|---|---|---|---|
| | Incidence rate ratio (95% CI) | P-value | Incidence rate ratio (95% CI) | P-value |
| **Pneumococcal or lobar pneumonia hospitalisation (n= 6 885 749§)** | 0.980 (0.976-0.984) | <0.001 | 0.985 (0.974-0.993) | 0.001 |
| **All-cause pneumonia hospitalisation (n=6 794 370§)** | 0.993 (0.992-0.994) | <0.001 | 0.991 (0.990-0.994) | <0.001 |

* adjusted by Indigenous status, age group, season, Index of Relative Socio-economic Advantage and Disadvantage (IRSAD) score, Accessibility or Remoteness Index of Australia (ARIA) category, birth weight, gestational age, maternal smoking during pregnancy, number of previous pregnancies, and previous hospitalisation for a range of high-risk medical comorbidities; †Any PCV – inclusive of both PCV7 and PCV13 § Number of observations for adjusted analysis (quarterly person-time)

#### G. Subgroup analyses – by age, rurality and Indigenous status

**Table 12: Crude and adjusted* incidence rate ratios of 7-valent pneumococcal conjugate vaccine-type (PCV7-type) invasive pneumococcal disease (IPD) in different sub-groups, by percent increase pneumococcal conjugate vaccine (PCV) coverage† among children under five years of age, New South Wales and Western Australia, 2009-2012**

| | Crude | | Adjusted | |
|---|---|---|---|---|
| | Incidence rate ratio (95% CI) | P-value | Incidence rate ratio (95% CI) | P-value |
| **Children < four months of age (n=300 118§)** | 0·983 (0·972-0·994) | 0·002 | 0·969 (0·939-1·001) | 0·056 |
| **Under-vaccinated children in urban areas (n=7 947 532§)** | 0·965 (0·961-0·970) | <0·001 | 0·966 (0·957-0·975) | <0·001 |
| **Under-vaccinated children in rural areas (n=632 239§)** | 0·965 (0·942-0·988) | 0·004 | 0·984 (0·946-1·003) | 0·438 |
| **Under-vaccinated Indigenous children (n=338 356§)** | 0·965 (0·941-0·990) | 0·006 | 0·981 (0·948-1·015) | 0·274 |

* adjusted by Indigenous status, season, Index of Relative Socio-economic Advantage and Disadvantage (IRSAD) score, Accessibility or Remoteness Index of Australia (ARIA) category, birth weight, gestational age, maternal smoking during pregnancy, number of previous pregnancies, and previous hospitalisation for a range of high-risk medical comorbidities; †Any PCV – inclusive of both PCV7 and PCV13; ‡Akaike information criterion § Number of observations for adjusted analysis (quarterly person-time)

**Table 13: Crude and adjusted* incidence rate ratios of all-cause pneumonia in different sub-groups, by percent increase in pneumococcal conjugate vaccine (PCV) coverage† among children under five years of age, New South Wales and Western Australia, 2001-2012**

| | Crude | | Adjusted | |
|---|---|---|---|---|
| | Incidence rate ratio (95% CI) | P-value | Incidence rate ratio (95% CI) | P-value |
| **Under-vaccinated children in urban areas (n=6 303 749)** | 0·992 (0·991-0·993) | <0·001 | 0·991 (0·990-0·993) | <0·001 |
| **Under-vaccinated children in rural areas (n=490 621)** | 0·998 (0·996-1·000) | 0·046 | 0·991 (0·986-0·997) | 0·002 |
| **Under-vaccinated Indigenous children (n=248 110)** | 0.986 (0.975-0.997) | 0.011 | 0.985 (0.958-1.012) | 0.274 |

* adjusted by Indigenous status, season, Index of Relative Socio-economic Advantage and Disadvantage (IRSAD) score, Accessibility or Remoteness Index of Australia (ARIA) category, birth weight, gestational age, maternal smoking during pregnancy, number of previous pregnancies, and previous hospitalisation for a range of high-risk medical comorbidities;

†Any PCV – inclusive of both PCV7 and PCV13; ‡Akaike information criterion § Number of observations for adjusted analysis (quarterly person-time)

#### H. Sensitivity analysis – association between coverage and indirection protection against invasive pneumococcal disease in un-vaccinated children

**Table 14: Crude and adjusted* incidence rate ratios of 7-valent pneumococcal conjugate vaccine-type (PCV7-type) invasive pneumococcal disease (IPD) among un-vaccinated children under five years, by percent increase in pneumococcal conjugate vaccine (PCV) coverage† among children under five years of age, New South Wales and Western Australia, 2009-2012**

| | Crude | | Adjusted* | |
|---|---|---|---|---|
| | Incidence rate ratio (95% CI) | P-value | Incidence rate ratio (95% CI) | P-value |
| **PCV7-type invasive pneumococcal disease (n= 8 579 771§)** | 0.967 (0.962-0.971) | <0.001 | 0.970 (0.962-0.978) | <0.001 |

**Table 15: Preventable fraction of PCV7-type IPD in un-vaccinated children under five years at deciles of pneumococcal conjugate vaccine (PCV) coverage, New South Wales and Western Australia, 2001-2012**

| Coverage (%) | Preventable fraction of PCV7-type IPD at deciles of PCV coverage |
|---|---|
| 10 | 26.0 (19.7-31.8) |
| 20 | 45.3 (35.5-53.5) |
| 30 | 59.5 (48.2-68.3) |
| 40 | 70.1 (58.4-78.4) |
| 50 | 77.8 (66.6-85.3) |
| 60 | 83.6 (73.2-90.0) |
| 70 | 87.9 (78.5-93.4) |
| 80 | 91.0 (82.7-95.3) |
| 90 | 93.4 (86.1-96.8) |
